# Prevalence of cardiovascular risk factors in osteoarthritis patients derived from primary care records: a systematic review of observational studies

**DOI:** 10.1101/2021.02.14.21251130

**Authors:** Xiaoyang Huang, Ross Wilkie, Mamas A Mamas, Dahai Yu

## Abstract

**Background:** People with osteoarthritis are at a high risk of cardiovascular disease (CVD). Detecting CVD risk factors in this high-risk population will help to improve CVD outcomes. Primary care electronic health records (EHRs) provide opportunities for the surveillance of CVD risk factors in the osteoarthritis population. This paper aimed to systematically review evidence of prevalence estimates of CVD risk factors in people with osteoarthritis derived from primary care EHRs.

**Methods:** Eight databases including MEDLINE were systematically searched to January 2019. Observational studies using primary care EHRs data to estimate the prevalence of six CVD risk factors in people with osteoarthritis were included. A narrative review was conducted to summarise study results.

**Results:** Six studies were identified. High heterogeneity between studies prevented the calculation of pooled estimates. One study reported the prevalence of smoking (12.5%); five reported hypertension (range: 19.7%-55.5%); four reported obesity (range: 34.4%-51.6%); two reported dyslipidaemia (6.0%, 13.3%); five reported diabetes (range: 5.2%-18.6%); and one reported chronic kidney disease (1.8%) in people with osteoarthritis. One study reported a higher prevalence of hypertension (Odds Ratio (OR) 1.25, 95% confidence interval (CI) 1.19-1.32), obesity (OR 2.44, 95%CI 2.33-2.55), dyslipidaemia (OR 1.24, 95%CI 1.14-1.35) and diabetes (1.11, 95%CI 1.02-1.22) in the osteoarthritis population compared with the matched non-osteoarthritis population.

**Conclusions:** From studies identified in this review that had used primary care EHRs, prevalence estimates of CVD risk factors were higher in people with osteoarthritis compared with those without. These estimates may provide baseline frequency of CVD risk factors in osteoarthritis patients in primary care, although this is limited by the small number of studies and high heterogeneity. Further studies of frequency, using primary care EHRs, will help to answer whether this data source can be used for evaluating approaches to manage CVD risk factors in osteoarthritis patients.

**Subject Area:** Primary care research

## Introduction

Health intelligence on the frequency of cardiovascular disease (CVD) and its risk factors identifies targets and populations for preventative strategies and allows evaluation of health outcomes, behaviours and interventions in populations and health care settings. Electronic health records (EHRs) represent a unique and still largely under-utilised source of data for health intelligence in high-risk primary care populations where strategies to reduce the burden of CVD are thought to have the greatest impact. The increasing availability of primary care EHRs provides opportunities for surveillance of risk factors for CVD (1–4). Proactive approaches to improving CVD outcomes and for recording patients’ data in primary care encourages the collection of information of the presence of CVD risk factors in healthcare populations (5–8). In the United Kingdom (UK), the Quality and Outcomes Framework (QOF) (7), an incentive payment programme which encourages general practitioners to record many conditions (e.g. smoking, hypertension, obesity, diabetes, dyslipidaemia and chronic kidney disease), improves the data capture of CVD risk factors as they are key QOF data items.

Osteoarthritis is extremely common in adults aged 60 and over and predicts CVD (9). Thirty-eight per cent of people with osteoarthritis have CVD compared to 9% of people without osteoarthritis (10). Traditional risk factors for CVD, including obesity, hypertension, dyslipidaemia, diabetes, are also associated with the development and progression of symptomatic osteoarthritis, potentially highlighting shared pathophysiological processes/pathways in their development (11–15). Primary care is the front-line setting of the identification and treatment of risk factors for CVD primary prevention in people with osteoarthritis (8). However, little is known about the co-occurrence of CVD risk factors in people with osteoarthritis recorded in primary care. A systematic review summarising evidence from mixed settings found a mean prevalence of dyslipidaemia in people with osteoarthritis of 30.2% (11). The review included only one study using primary care EHRs that reported a markedly lower estimate (13.3%) (16). Understanding the prevalence of CVD risk factors with OA recorded in primary care is important to inform best practice. This systematic review aimed to identify and evaluate studies to estimate the prevalence of CVD risk factors including smoking, obesity, hypertension, dyslipidaemia, diabetes and chronic kidney disease in people with osteoarthritis using primary care EHRs.

## Methods

This systematic review was conducted with reference to the PRISMA guidelines for reporting systematic reviews (17). The protocol of this systematic review was registered on PROSPERO (CRD42018088405).

### Search strategy

One reviewer (XH) identified studies by searching eight electronic databases, including MEDLINE, EMBASE, PsycINFO, COCHRANE LIBRARY, PUBMED, CINAHL, AMED and WEB OF SCIENCE from their inception to November 2017 and updated the search in January 2019. The review also scanned reference lists of all included papers and relevant studies for additional eligible studies. Details of the search strategy for MEDLINE is showed in the supplementary materials.

### Study selection

Studies were considered eligible if they were cross-sectional, cohort or case-control studies and estimated the prevalence of cardiovascular risk factors, including smoking, hypertension, obesity, dyslipidaemia, diabetes or chronic kidney disease, in people with osteoarthritis using data from primary care EHRs. No restriction was imposed on participants’ age, gender, ethnicity, or their severity or localisation of osteoarthritis. Studies were included if they identified people with osteoarthritis as those with a recorded osteoarthritis diagnosis or osteoarthritis-related joint pain. Smoking was defined as a record of current smoking status; hypertension as a record of hypertension diagnosis, high systolic blood pressure (≥ 140 mm Hg)/diastolic blood pressure (≥ 90 mm Hg) or being prescribed antihypertensive drugs; obesity as a record of obesity, or high body mass index (BMI) (≥ 30kg/m^2^)/waist circumference (men: ≥ 94cm; women: ≥ 80cm)/waist-hip ratio (men: ≥ 0.9; women: ≥ 0.85); dyslipidaemia as a record of dyslipidaemia diagnosis, high level of serum total cholesterol (≥ 5 mmol/L)/LDL-C (≥ 3 mmol/L)/triglyceride (≥ 1.7 mmol/L), low HDL-C (men: <1 mmol/L; women: <1.2 mmol/L), or being prescribed lipid-lowering drugs; diabetes as a record of type 1/type 2 diabetes diagnosis, high fasting blood glucose (≥ 7.0 mmol/L)/2-hour blood glucose (≥ 11.1 mmol/L)/HbA1c (≥ 6.5%), or being prescribed antidiabetic drugs/insulin; chronic kidney disease as a record of chronic kidney disease diagnosis, high urinary albumin: creatinine ratio (≥ 3mg/mmol), low estimated glomerular filtration rate (<60 ml/min/1.73 m^2^), or engagement of renal replacement therapy.

Studies using general osteoarthritis patients were considered eligible. To avoid overestimating the prevalence of CVD risk factors in osteoarthritis, studies were excluded if they used a subgroup of osteoarthritis population with a particular high CVD risk (i.e. studies only have osteoarthritis patients with previous or comorbid CVD, smoking, hypertension, obesity, dyslipidaemia, diabetes or chronic kidney disease). Studies not published in English were also excluded.

Three reviewers (XH, DY and RW) independently reviewed the titles, abstracts and full texts of citations identified through the search of electronic databases. Any disagreements were resolved through discussion to consensus.

### Data extraction

One reviewer (XH) independently extracted data about key information (e.g. study design, country, age and gender distribution in the study population, characteristics of the primary care database used, the definition of osteoarthritis, definition of CVD risk factors, raw number and prevalence estimates) from the full text of included studies and another two reviewers (DY and RW) verified the extracted data.

### Risk of bias assessment

Three reviewers (XH, DY and RW) independently examined the risk of bias in included studies using the Quality in Prognosis Studies (QUIPS) tool and resolved disagreements by consensus (18). The QUIPS tool offers criteria for assessing six important domains of bias, including participation, attrition, measurement of exposure, measurement of outcomes, confounding account and statistical analysis. The overall risk of bias in a domain was low where reviewers rated “yes’ or “not applicable”, moderate where rated “partial” or “unsure” and high where rated “no” to the summary statement of the domain.

### Data analysis

Meta-analyses were attempted to provide pooled prevalence estimates for each CVD risk factor in the overall osteoarthritis population using a random-effects model (19,20). However, the extent of heterogeneity (e.g. I^2^=99.9% for the pooled hypertension prevalence) between studies indicate that pooling estimates were not appropriate. The small number of identified studies prevented subgroup analyses to assess if the prevalence estimate was influenced by age, gender or location of osteoarthritis.

Prevalence estimates of each CVD risk factor in people with and without osteoarthritis from included studies were presented in tables and texts. Raw counts where available were used to calculate the odds ratio (OR) and confidence interval (CI) for each CVD risk factor in people with osteoarthritis. A narrative review was performed to explore potential sources of heterogeneity between included studies for age, gender, the length of prevalence period, characteristics of the primary care database used, the definition of osteoarthritis, definition of CVD risk factors, and inclusion of potential confounders.

## Results

### Search results

The systematic search identified 23,076 articles after the removal of 7,774 duplicates (Figure 1). Following the title review, 509 articles were included in the abstract review. Of these, 21 papers were included for full-text review, of which 6 papers met the inclusion criteria; no study was reported more than once and six studies were considered in the analysis (16,21–25).

### Data quality

All six studies were of good methodological quality. Reviewers agreed the risk of selection, measurement and non-response bias was low. The greatest risk of bias was due to confounding. Two studies were rated as having a moderate risk of bias due to a failure to account for potential confounders when comparing prevalence estimates among people with and without osteoarthritis (16,23).

### Study findings

The six identified studies estimated the prevalence of at least one of the six CVD risk factors in people with osteoarthritis (16,21–25). Three also reported prevalence in people without osteoarthritis and compared these to those with OA (16,23,24) (Table 1). Four were cohort studies (22–25) and two were cross-sectional studies (16,21). The primary care EHR data included in the studies came from databases of general practice consultation records in British Columbia, Canada (n=1) (24), Catalonia, Spain (n=2) (22,23), and Tayside, Scotland (n=1) (25); a national primary care database in the Netherlands (16), and computerised health records from four general practices within Mexico City, Mexico (21). The population with osteoarthritis in each study was older than those without osteoarthritis (mean age ranged from 58 to 70 years in the osteoarthritis population compared to 51 to 61 years in the non-osteoarthritis population). The proportion of females was higher in the osteoarthritis population than the non-osteoarthritis population (range 59%-74% vs. 50%-59%). Only one study excluded individuals with a history of CVD prior to the osteoarthritis diagnosis (26).

**Table 1.** Characteristics of included studies.

| **Study author, publication year** | **Study design** | **Data source** | **Primary care EHRs** | **Identification of OA** | **Sample size** | **Mean age (years)** | **Females (%)** | **CVD (%)** |
| --- | --- | --- | --- | --- | --- | --- | --- | --- |
| Doubov a et al, 2015 | Cross-sectional | Mexican Institute of Social Security electronic health records | Regional EHRs containing around 585,535 persons' medical information from four primary care practices in Mexico City, Mexico | At least one visit with an ICD-10 code for knee and hip OA (M160-M17X) | OA: 8,991 | OA: 60 | OA: 69% | OA:4.5% |
| Leyland et al, 2016 | Cohort | Information System for Development of Primary Care Research (SIDIAP) | A regional database containing over 5.5 million persons' medical information from over 270 primary care practices in Catalonia, Spain | At least one visit with an ICD-10 code for knee OA (M17) | OA: 97,677 | OA: 68 | OA: 66% | Not reported |
| Nielen et al, 2012 | Cross-sectional | Netherlands Information Network of General Practice (LINH) | A national database containing around 360,000 persons' medical information from 96 primary care practices in the Netherlands | At least one visit with a ICPC-1 code for knee and hip OA (L89 and/or L90) | OA: 4,040<br>Non-OA: 158,439 | OA: 70;<br>Non-OA: 51 | OA: 69%<br>Non-OA: 50% | OA:5.8%<br>Non-OA: 2.0% |
| Prieto-Alhambra et al, 2014 | Cohort | SIDIAP | A regional database containing over 5.5 million persons' medical information from over 270 primary care practices in Catalonia, Spain | At least one visit with an ICD-10 code for knee, hip and hand OA (M17, M16, M15.1, M15.2, M18) | Knee OA: 96,222 ;<br>Hip OA: 30,350 ;<br>Hand OA: 37,590<br>Non-OA: 2,955, 289 | Knee OA: 67; ;<br>Hip OA: 69; ;<br>Hand OA: 64; ;<br>Non-OA: 61 | Knee OA: 64% ;<br>Hip OA: 58% ;<br>Hand OA: 74% ;<br>Non-OA: 50% | Knee OA: 3.3% ;<br>Hip OA: 4.0% ;<br>Hand OA: 3.5% ;<br>Non-OA: Not reported |
| Rahman et al, 2013 | Cohort | British Columbia Ministry of Health administrative database | A regional database containing 600,000 randomly selected residents' health information in British Columbia, Canada | At least two visits with an ICD-9 (715) or ICD-10 code for OA (M15–M19) | OA: 12,745<br>Non-OA: 36,886 | OA: 58; ;<br>Non-OA: 58 | OA: 60% ;<br>Non-OA: 59% | OA: 0% ;<br>Non-OA: 0% |
| Sheng et al, 2012 | Cohort | Medicines Monitoring (MEMO) Unit record-linked databases | A regional database containing resident's health information in Tayside, Scotland | At least one visit with an ICD-9 (715) or ICD-10 code for OA (M15-19, or M47) | OA: 1,269 | OA: 69 | OA: 59% | OA: 1.6% |

OA, osteoarthritis;
CVD: cardiovascular disease
EHRs, electronic health records
ICD-9, International Statistical Classification of Diseases and Related Health Problems 9<sup>th</sup> version;
ICD-10, International Statistical Classification of Diseases and Related Health Problems 10<sup>th</sup> version;
ICPC-1, International Classification of Primary Care 1<sup>ST</sup> version

Prevalence estimates of CVD risk factors in people with osteoarthritis

Five studies provided prevalence estimates of hypertension in osteoarthritis with a range from 19.7% to 55.5% (16,21,23–25).

Five studies reported prevalence estimates of diabetes in osteoarthritis with a range from 5.2% to 18.6% (16,21,23–25).

Four studies that assessed the obesity prevalence in osteoarthritis showed a range between 34.4% and 51.6% (21–24).

Two studies estimated the prevalence of dyslipidaemia in osteoarthritis (16,24). One estimate (6.0%) was from an osteoarthritis population aged 20 and over without a history of CVD in a study using an administrative database in British Columbia, Canada (24). Another estimate (13.3%) was from people with knee or hip osteoarthritis aged over 30 years provided by a study using a national primary care database in the Netherlands (16).

Only one of the studies reported the prevalence of chronic kidney disease (1.8% in people with knee or hip osteoarthritis who aged 20 and over from four practices in Mexico City) (21) and one study estimated the prevalence of smoking (12.5% in knee osteoarthritis patients aged over 40 years with no history of knee replacement over a six-year) (22).

### Association between osteoarthritis and CVD risk factors

Three of the six studies provided estimates of the prevalence of CVD risk factors in people with and without osteoarthritis (16,23,24)(Table 2). All three studies reported a positive association between osteoarthritis and CVD risk factors; this was reported for hypertension (16,24), obesity (23,24), dyslipidaemia (16,24), and type 2 diabetes (24). Only one study accounted for confounders; the odds ratios (ORs) calculated based on the age-gender-standardised prevalence estimates showed that osteoarthritis was significantly associated with a higher prevalence of hypertension (OR 1.25, 95%CI 1.19 to 1.32), obesity (OR 2.44, 95%CI 2.33 to2.55), dyslipidaemia (OR 1.24, 95%CI 1.14 to 1.35) and type 2 diabetes (1.11, 95%CI 1.02 to 1.22) (24).

**Table 2.**
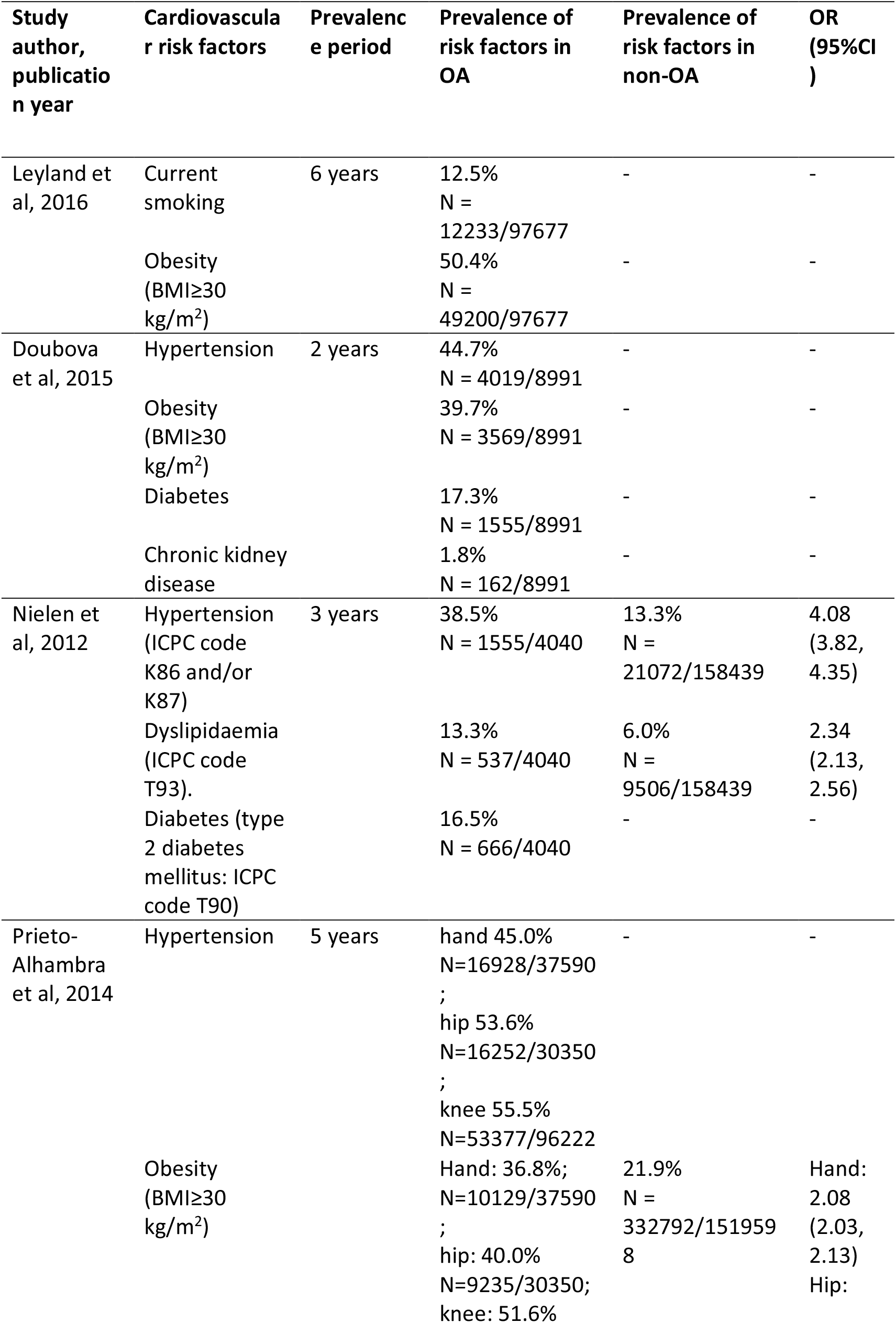
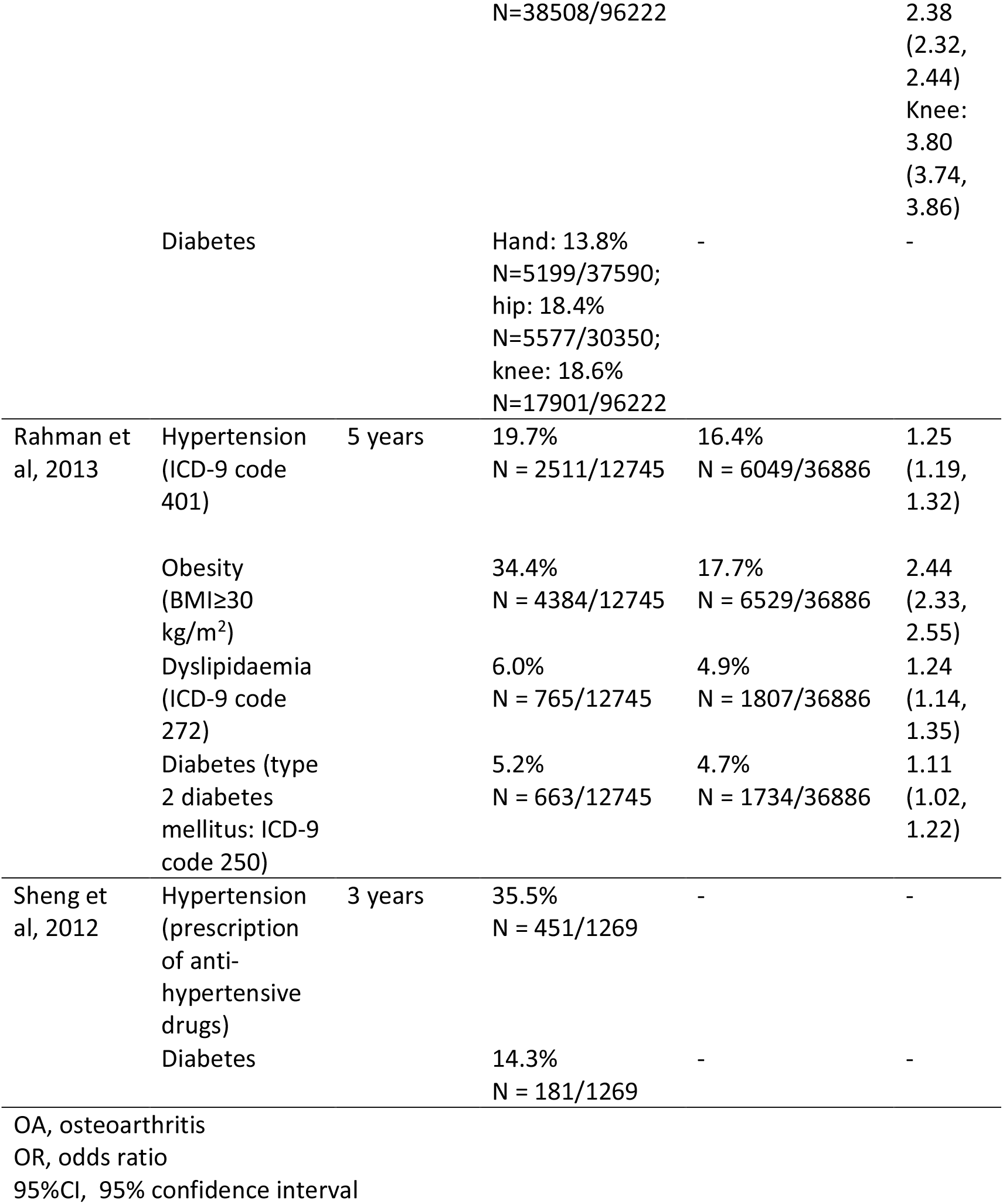
Findings from included studies.

### Narrative review of potential sources of heterogeneity

#### Heterogeneity by age and gender

None of the studies reported age- or gender-stratified prevalence of any CVD risk factors in the osteoarthritis population. The mean age and gender distribution of the osteoarthritis population varied markedly between studies reporting prevalence estimates of hypertension, obesity, hypertension or diabetes. Regarding dyslipidaemia, the study in which the osteoarthritis population had a higher mean age (70 years) and proportion of females (69%) reported a higher prevalence estimate (13.3% vs. 6.0%) compared with another study (mean age: 58 years; the proportion of females: 60%) (16,24).

#### Heterogeneity by definition and locality of osteoarthritis

The prevalence of CVD risk factors varied between osteoarthritis populations with different joint affected but this variation might due to differences in the population. Four studies identified knee and/or hip osteoarthritis (16,21,22); two identified generalised osteoarthritis (24,25); and one identified hand osteoarthritis (23). Among the four studies reporting obesity, knee and/or hip osteoarthritis populations had the highest prevalence (39.7%-51.5%); the hand osteoarthritis population reported a lower estimate (36.8%); and the generalised osteoarthritis population the lowest (34.4%) (21–24). However, the difference in the prevalence is likely to be affected by differences in the age and gender distribution of the osteoarthritis sample. Between the two studies reporting dyslipidaemia, the knee and/or hip osteoarthritis population in one study showed a higher prevalence estimate (13.3% vs. 6.0%) than the generalised osteoarthritis population in another (16,24). Whether joint location explained the variation in the prevalence of hypertension or diabetes between studies was unclear. Only one study reported joint-specific prevalence of risk factors (18); based on data from the SIDIAP database, people with knee osteoarthritis had the highest prevalence estimate of hypertension (55.5%), obesity (51.6%) and type 2 diabetes (18.6%); hip osteoarthritis had a slightly lower figure (53.6%, 40.0% and 18.4%); and hand osteoarthritis with the lowest estimate (45.0%, 36.8% and 13.8%).

The most commonly used coding standard for osteoarthritis identification in the included studies (n=5) was the International Statistical Classification 9^th^ (ICD-9) or 10^th^ version (ICD-10) but the choice of codes was unique in each study (Table 1). The broadest definition of osteoarthritis (ICD-9 code 715 and ICD-10 code M15-19, or M47) among studies using ICD codes was adopted by Sheng et al (2012) who reported a prevalence rate of hypertension as 35.5% and diabetes as 14.3% in osteoarthritis (25). Only one study used International Classification of Primary Care 1^st^ version (ICPC-1) to identify knee and hip osteoarthritis cases and estimated the prevalence of hypertension as 38.5%, dyslipidaemia as 13.3% and diabetes as 16.5% in osteoarthritis (16).

#### Heterogeneity by definition of CVD risk factors

Only two studies reported codes used to identify CVD risk factors and there were inconsistencies between them (16,24). Nielen’s (2012) study which used the ICPC-1 code reported a higher prevalence of CVD risk factors (hypertension 38.5%, dyslipidaemia as 13.3%, and type 2 diabetes as 16.5%) in osteoarthritis patients compared with Rahman et al’s (2013) study which used ICD-9 code (hypertension 19.7%, dyslipidaemia 6.0%, and type 2 diabetes 5.2%).

## Discussion

### Summary of evidence

This review summarised the evidence of the prevalence of CVD risk factors in people with osteoarthritis from primary care EHRs to obtain a baseline prevalence estimate of CVD risk factors in this high-risk population with osteoarthritis. Prevalence estimates of hypertension, obesity, dyslipidaemia and diabetes in people with osteoarthritis derived from primary care EHRs varied considerably between studies. Evidence on prevalence estimates of smoking and chronic kidney disease in osteoarthritis from general practice records are limited. Osteoarthritis is significantly associated with a higher prevalence of hypertension, obesity, dyslipidaemia and type 2 diabetes. The robustness of these associations is unclear because of the small number of reviewed studies, heterogeneous populations studied and disease definitions used, the high potential for bias and unmeasured confounders.

Comparison of study results was challenging because of the differences in population characteristics. Although it was also not clear whether variations in age and gender distribution between osteoarthritis populations affected the reported prevalence estimates of CVD risk factors from the evidence base identified by this systematic review, as older age and female gender may confound the observed association between osteoarthritis and CVD risk factors. The risk of CVD, as well as risk factors including dyslipidaemia, hypertension and diabetes, are higher in older age groups (26). Thus, an older population was likely to include more cases with CVD risk factors compared with a younger population. This might contribute to our findings that the association between osteoarthritis and cardiovascular risk factors observed from age- and gender-matched populations with and without osteoarthritis (24) was smaller than that from unmatched populations (16,23).

The prevalence of some CVD risk factors in people with osteoarthritis derived from primary care EHRs is close to that from other data sources. Data from 168 outpatients with osteoarthritis found a slightly lower prevalence of obesity at 30% compared with estimates from the reviewed studies (range 34.4%-51.6%) (27). A population-based survey including 24.3 million adults with osteoarthritis aged 35 and over reported prevalence estimates of hypertension (40%) and diabetes (11%) in OA within the prevalence range (hypertension: 19.7% to 55.5%, diabetes: 5.2% to 18.6%) identified by this systematic review (13). However, prevalence estimates of current smoking (20%) and dyslipidaemia (32%) in osteoarthritis from the survey were markedly higher than those derived from primary care EHRs used in the reviewed studies (current smoking: 12.2%, dyslipidaemia: 13.3%) (16,22,24). This suggests that smoking and dyslipidaemia in people who consult general practices for osteoarthritis may not be robustly identified or coded in primary care EHRs, and as a result may be undertreated in this population. The prevalence of chronic kidney disease in osteoarthritis (0.8%) from the survey was similar than that from the included study reporting related figure (1.8%) (13). However, the different measurement of cardiovascular risk factors (e.g. laboratory data vs. doctor’s diagnosis) and the lack of information on diagnostic criteria used in each study make it hard to confirm the disparity in prevalence between studies.

The review findings suggest that people consulting primary care providers with osteoarthritis had higher prevalence rates of hypertension, obesity, dyslipidaemia and type 2 diabetes than those without osteoarthritis. These findings are consistent with what has been shown in other settings (11,13). Baudart et al.’s systematic review reported a higher pooled estimate of dyslipidaemia as 30.2% in people with osteoarthritis than the 8.0% in those without osteoarthritis (11). There is evidence from a population-based survey which suggested that the US population aged over 35 years with osteoarthritis had higher prevalence estimates of hypertension (40% vs.25%), dyslipidaemia (32% vs.24%) as well as diabetes (11% vs. 6%) than the general population without osteoarthritis (13).

### Strengths and limitations

This review provides a synthesis of evidence on the prevalence of CVD risk factors in osteoarthritis derived from primary care EHRs. The focus on data from primary care EHRs allowed the identification of studies with large sample size, a good reflection of routine practice and an internal reference group for comparing people with and without osteoarthritis. The studies identified included data from representative samples of the general population.

There were limitations in this systematic review. The small number of reviewed studies and the high heterogeneity in age and gender distribution, definition of osteoarthritis and CVD risk factors made it impossible to conduct a statistical combination of prevalence estimates. However, a narrative review of possible reasons of variation which has been performed here is useful to avoid potentially biased results generated by pooling estimated from studies with high heterogeneity (28). This review does not provide insight into whether there are differences in CVD risk factor profile depending on the severity/period of the osteoarthritis. The severity/period of the osteoarthritis might affect the prevalence estimate of CVD risk factors because chronic inflammation included in osteoarthritis aetiology has often been proposed to explain the link between osteoarthritis and CVD (29). An additional limitation to consider is the under-reported CVD risk factors in EHRs. Data from EHRs might under-detect some risk factors (such as those not included in quality incentive schemes), resulting in an underestimate of actual prevalence (30). In the reviewed studies, CVD risk factors more frequently registered in primary care EHRs might be conditioned by the fact that they are of particular interest, such as hypertension, obesity and diabetes in many quality incentive schemes. There was a lack of adjustment for confounders, such as age and gender, which may have a substantial impact on the observed association between osteoarthritis and CVD risk factors in the reviewed studies. Moreover, the investigation of CVD risk factors of osteoarthritis patients with established CVD could be more frequently and the prevalence estimate might be overestimated but there was only one study excluded these patients. Finally, this review only provided binary estimates about the presence/absence of a CVD risk factor rather than considering the actual measurements of cholesterol, blood pressure, BMI etc, that are as important for understanding CVD risk as the presence of the risk factors themselves.

## Conclusions

This systematic review found substantial variation in prevalence estimates of CVD risk factors in osteoarthritis using data from primary care EHRs. People who have consulted GPs for osteoarthritis are more likely to have a higher prevalence of hypertension, obesity, dyslipidaemia and diabetes compared with those without osteoarthritis. However, the small number of studies and high heterogeneity identified across the studies fails to indicate the expected prevalence level of CVD risk factors in people with osteoarthritis for comparison with populations that do not have osteoarthritis. This suggests that further studies using large-scale and good representative primary care EHR data is required to identify a baseline prevalence to then allow evaluation of whether these risk factors are being identified and if interventions focusing on prevention are successful. Such studies should consider using similar methods, including comparable populations and methods identifying risk factors and conditions. Future work on the standardisation of disease definition applied to primary care EHRs on an international scale may improve the comparison of disease frequency between studies and facilitate more precise estimates. Furthermore, future work should study whether different anatomical sites of osteoarthritis are associated with different CVD risk factor profiles and examine whether there is a relationship between CVD risk factors and osteoarthritis severity based on clinical parameters. While the association between osteoarthritis and CVD risk factors is not robust, prevalence estimates derived from primary care EHRs suggest that there is a need to intensively detect and treat CVD risk factors in osteoarthritis patients in clinical practice.

## Data Availability

This review was pre-registered at PROSPERO (CRD42018088405).

https://www.crd.york.ac.uk/prospero/display_record.php?ID=CRD42018088405

## Acknowledgements

The authors would like to thank Dr Nadia Corp from the systematic review team in School of Primary, Community and Social Care at Keele University for revising the search strategy used in this systematic review.

---

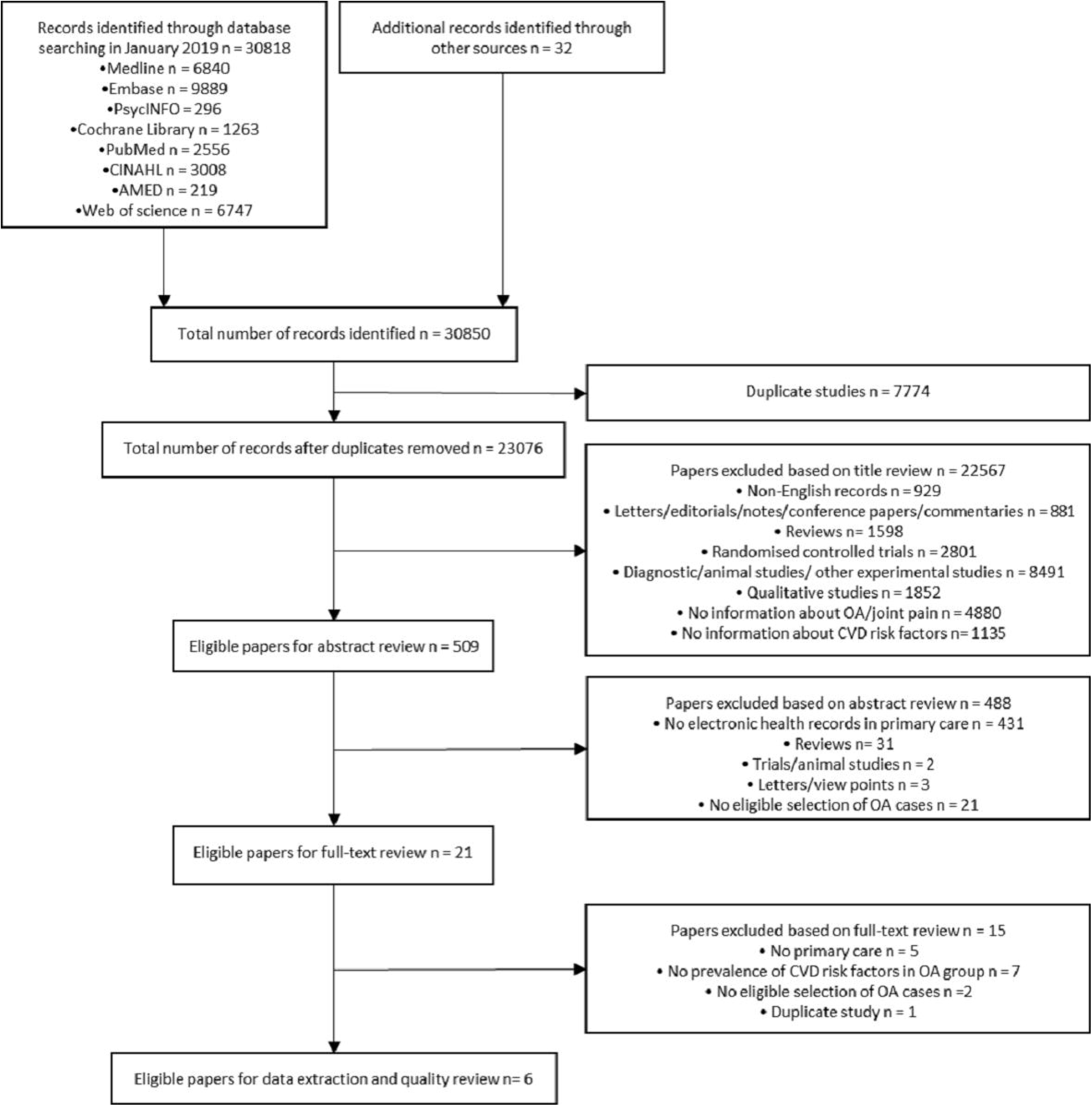

