## supplementary materials for "Prevalence of cardiovascular risk factors in osteoarthritis patients derived from primary care records: a systematic review of observational studies"

**Table S1. Search strategy for MEDLINE**

| Medline (OvidSP) (Ovid MEDLINE(R) Epub Ahead of Print, In-Process & Other Non-Indexed Citations, Ovid MEDLINE(R) Daily and Ovid MEDLINE(R) 1946 to January 15, 2019) | | |
| --- | --- | --- |
|  | Searches | Results |
| 1 | exp Osteoarthritis/ | 57392 |
| 2 | osteoarthr$.tw. | 62819 |
| 3 | OA.tw. | 30410 |
| 4 | (degenerative adj (arthritis or joint or joints)).tw. | 4031 |
| 5 | arthrosis.tw. | 5163 |
| 6 | (knee$ adj3 (pain or painful)).tw. | 9846 |
| 7 | (hip$ adj3 (pain or painful)).tw. | 5761 |
| 8 | (hand$ adj3 (pain or painful)).tw. | 2131 |
| 9 | (joint$ adj3 (pain or painful)).tw. | 11114 |
| 10 | (finger$ adj3 (pain or painful)).tw. | 573 |
| 11 | (thumb$ adj3 (pain or painful)).tw. | 231 |
| 12 | (shoulder$ adj3 (pain or painful)).tw. | 8497 |
| 13 | ((foot or feet) adj3 (pain or painful)).tw. | 2424 |
| 14 | (ankle$ adj3 (pain or painful)).tw. | 1624 |
| 15 | or/1-14 | 130108 |
| 16 | exp Cardiovascular Diseases/ | 2242925 |
| 17 | exp Risk/ | 1104635 |
| 18 | 16 and 17 | 284119 |
| 19 | ((cardiovascular$ or cardio-vascular$ or CVD or CV or coronary or CHD or "heart disease$" or cerebrovascular$ or "heart failure$" or HF or stroke$ or isch?emia$ or "heart decompensation$" or "myocardial infarct$" or MI or angina) adj3 risk).tw. | 168896 |
| 20 | 18 or 19 | 372619 |
| 21 | Smoking/ | 134655 |
| 22 | Tobacco/ | 29146 |
| 23 | (Tobacco$ or smoking or smoke$ or cigarette$ or cigar$).tw. | 307855 |
| 24 | exp Dyslipidemias/ | 75560 |
| 25 | dyslipid?emia$.tw. | 29232 |
| 26 | Dyslipoprotein?emia$.tw. | 981 |
| 27 | (hyperlipid?emia$ or hyper-lipid?emia$).tw. | 24627 |
| 28 | (hyperlipoprotein?emia$ or hyper-lipoprotein?emia$).tw. | 4285 |
| 29 | (hypercholesterol?emi$ or hyper-cholesterol?emi$).tw. | 32572 |
| 30 | (hypertriglycerid?emia$ or hyper-triglycerid?emi$).tw. | 11953 |
| 31 | ((elevat$ or high$ or increas$) adj3 (cholesterol or TC or "low-density lipoprotein" or LDL or LDL-C or triglyceride$ or TG)).tw. | 92710 |
| 32 | ((reduc$ or low$ or decreas$) adj3 ("high density lipoprotein" or HDL or HDL-C)).tw. | 25136 |
| 33 | exp Hypolipidemic Agents/ | 132892 |
| 34 | exp Hydroxymethylglutaryl-CoA Reductase Inhibitors/ | 37415 |
| 35 | (antilipid$ or anti-lipid$).tw. | 1035 |
| 36 | ("Hydroxymethylglutaryl-CoA Reductase Inhibit$" or "HMG-COA reductase inhibit$" or Statin$).tw. | 41065 |
| 37 | exp Diabetes Mellitus/ | 395207 |
| 38 | exp Diabetes Complications/ | 123985 |
| 39 | Blood Glucose/ | 155162 |
| 40 | Hemoglobin A, Glycosylated/ | 31857 |
| 41 | Metabolic Syndrome X/ | 28457 |
| 42 | (diabete$ or diabetic$ or DM or T1D or T1DM or T2D or T2DM).tw. | 589733 |
| 43 | "metabolic syndrome$".tw. | 45104 |
| 44 | ((elevat$ or high$ or increas$) adj3 ("blood glucose" or "blood sugar" or HbA1c)).tw. | 14150 |
| 45 | exp Agents, Hypoglycemic/ | 234048 |
| 46 | exp Sulfonylurea Compounds/ | 18638 |
| 47 | exp Biguanides/ | 24468 |
| 48 | Sodium-Glucose Transporter 2/ | 1414 |
| 49 | alpha-Glucosidases/ | 4221 |
| 50 | Glucagon-Like Peptide 1/ | 7100 |
| 51 | Thiazolidinediones/ | 11105 |
| 52 | exp Amylin Receptor Agonists/ | 2320 |
| 53 | (antidiabet$ or anti-diabet$).tw. | 20824 |
| 54 | insulin.tw. | 333831 |
| 55 | ("Sodium glucose co-transporter 2" or "Sodium glucose transporter 2").tw. | 759 |
| 56 | "Sulfonylurea Compound$".tw. | 155 |
| 57 | Biguanide$.tw. | 2708 |
| 58 | "alpha-glucosidase inhibit$".tw. | 2522 |
| 59 | "glucagon-like peptide-1".tw. | 9400 |
| 60 | thiazolidinedione$.tw. | 5481 |
| 61 | "amylin analog$".tw. | 131 |
| 62 | exp Obesity/ | 192968 |
| 63 | Body Mass Index/ | 114794 |
| 64 | exp Body weight/ | 432419 |
| 65 | exp Body Fat Distribution/ | 12418 |
| 66 | weight gain/ | 29398 |
| 67 | exp Waist Circumference/ | 9022 |
| 68 | Waist-Hip Ratio/ | 3813 |
| 69 | exp Adipose Tissue/ | 90570 |
| 70 | (adipos$ or obes$).tw. | 327945 |
| 71 | ("body mass ind$" or "body mass" or BMI).tw. | 239950 |
| 72 | "weight gain".tw. | 56289 |
| 73 | ("waist circumference$" or "waist-hip ratio").tw. | 26468 |
| 74 | (fat or "body fat distribution" or "fat overload syndrom$").tw. | 237221 |
| 75 | (overeat$ or over-eat$ or overfeed$ or over-feed$).tw. | 4617 |
| 76 | exp Renal insufficiency, chronic/ | 105787 |
| 77 | ((endstage or end stage or established or chronic or progressive) adj1 (renal or kidney) adj1 (failure or disease$ or insufficien$)).tw. | 102442 |
| 78 | exp Dialysis/ | 23279 |
| 79 | Dialysis.tw. | 100458 |
| 80 | (ESKD or ESRD or ESRF).tw. | 16181 |
| 81 | (CKD or CKF or CKI or CRD or CRF or CRI).tw. | 44459 |
| 82 | (h?emodialysis or h?emofiltration or h?emodiafiltration).tw. | 75510 |
| 83 | (predialysis or pre-dialysis).tw. | 4728 |
| 84 | exp Hypertension/ | 242941 |
| 85 | hypertens$.tw. | 399868 |
| 86 | ((elevat$ or high$ or increas$) adj3 BP).tw. | 13258 |
| 87 | ((elevat$ or high$ or increas$) adj3 systolic).tw. | 16293 |
| 88 | ((elevat$ or high$ or increas$) adj3 SBP).tw. | 4150 |
| 89 | ((elevat$ or high$ or increas$) adj3 diastolic).tw. | 10756 |
| 90 | ((elevat$ or high$ or increas$) adj3 DBP).tw. | 1939 |
| 91 | ((elevat$ or high$ or increas$) adj3 "blood pressur$").tw. | 56736 |
| 92 | exp Antihypertensive Agents/ | 246285 |
| 93 | (antihypertensi$ or anti-hypertensi$).tw. | 49675 |
| 94 | exp Angiotensin-Converting Enzyme Inhibitors/ | 42141 |
| 95 | exp Angiotensin Receptor Antagonists/ | 21383 |
| 96 | exp Adrenergic beta-Antagonists/ | 81654 |
| 97 | exp Adrenergic Antagonists/ | 120653 |
| 98 | exp Thiazides/ | 15169 |
| 99 | exp sodium chloride symporter inhibitors/ | 13943 |
| 100 | exp sodium potassium chloride symporter inhibitors/ | 13471 |
| 101 | exp Diuretics/ | 77509 |
| 102 | (angiotensin adj3 (receptor antagon$ or receptor block$)).tw. | 13203 |
| 103 | ARB$.tw. | 75511 |
| 104 | (beta adj3 (adrenergic$ or antagonist$ or block$ or receptor$)).tw. | 111311 |
| 105 | (alpha adj3 (adrenergic$ or antagonist$ or block$ or receptor$)).tw. | 83588 |
| 106 | ((angiotensin or adrenergic$) adj3 antagonist$).tw. | 11804 |
| 107 | Thiazide$.tw. | 5289 |
| 108 | diuretic$.tw. | 36153 |
| 109 | "angiotensin converting enzyme inhibit$".tw. | 19269 |
| 110 | (ACE adj2 inhibit$).tw. | 18968 |
| 111 | ACEI$.tw. | 4114 |
| 112 | exp Calcium Channel Blockers/ | 78983 |
| 113 | (calcium adj3 (antagonist$ or block$ or inhibit$)).tw. | 43769 |
| 114 | CCB$.tw. | 3632 |
| 115 | or/20-114 | 3325979 |
| 116 | Epidemiologic Studies/ | 7849 |
| 117 | exp Case-Control Studies/ | 965570 |
| 118 | exp Cohort Studies/ | 1816093 |
| 119 | Cross-Sectional Studies/ | 283967 |
| 120 | epidemiolog$.tw. | 342858 |
| 121 | ("case control$" or case-control$).tw. | 115935 |
| 122 | Cohort$.tw. | 494647 |
| 123 | ("cross sectional" or cross-sectional).tw. | 298451 |
| 124 | ("follow up" or follow-up).tw. | 867827 |
| 125 | longitudinal.tw. | 215899 |
| 126 | retrospective$.tw. | 637977 |
| 127 | prospective$.tw. | 629671 |
| 128 | (observ$ adj3 (study or studies)).tw. | 164062 |
| 129 | or/116-128 | 3629310 |
| 130 | 15 and 115 and 129 | 6912 |
| 131 | exp animals/ not humans/ | 4538799 |
| 132 | 130 not 131 | 6840 |
